# Emergence and Spread of the SARS-CoV-2 Variant of Concern Delta Across Different Brazilian Regions

**DOI:** 10.1101/2021.11.25.21266251

**Authors:** Ighor Arantes, Felipe Gomes Naveca, Tiago Gräf, COVID-19 Fiocruz Genomic Surveillance Network, Fábio Miyajima, Helisson Faoro, Gabriel Luz Wallau, Edson Delatorre, Luciana Reis Appolinario, Elisa Cavalcante Pereira, Taina Moreira Martins Venas, Alice Sampaio Rocha, Renata Serrano Lopes, Marilda Mendonça Siqueira, Gonzalo Bello, Paola Cristina Resende

**Author notes:** Authors and affiliations detailed in Appendix 01.

## Abstract

The SARS-CoV-2 Variant of Concern (VOC) Delta was first detected in India in October 2020. The first imported cases of the Delta variant in Brazil were identified in April 2021 in the Southern region, followed by more cases in different country regions during the following months. By early September 2021, Delta was already the dominant variant in the Southeastern (87%), Southern (73%), and Northeastern (52%) Brazilian regions. This work aimed to understand the spatiotemporal dissemination dynamics of Delta in Brazil. To this end, we employed a combination of Maximum Likelihood (ML) and Bayesian methods to reconstruct the evolutionary relationship of 2,264 of VOC Delta complete genomes (482 from this study) recovered across 21 out of 27 Brazilian federal units. Our phylogeographic analyses identified three major transmission clusters of Delta in Brazil. The clade BR-I (*n* = 1,560) arose in Rio de Janeiro in late April 2021 and was the major cluster behind the dissemination of the VOC Delta in the Southeastern, Northeastern, Northern, and Central-Western regions. The clade BR-II (*n* = 207) arose in the Paraná state in late April 2021 and aggregated the largest fraction of sampled genomes from the Southern region. Lastly, the clade BR-III emerged in the São Paulo state in early June 2021 and remained mostly restricted to this state. In the rapid turnover of viral variants characteristic of the SARS-CoV-2 pandemic, Brazilian regions seem to occupy different stages of an increasing prevalence of the VOC Delta in their epidemic profiles. This process demands continuous genomic and epidemiological surveillance toward identifying and mitigating new introductions, limiting their dissemination, and preventing the establishment of more significant outbreaks in a population already heavily affected by the COVID-19 pandemic.

## INTRODUCTION

The severe acute respiratory syndrome coronavirus 2 (SARS-CoV-2) variant of concern (VOC) Delta (B.1.617.2+AY.*/ G/ 478K.V1/ 21A) was first described in India in October 2020. By April 2021, it was designated by the World Health Organization (WHO) as a Variant of Interest (VOI) and on May 11, 2021, was formally recognized as a VOC (WHO, 2021). Delta possesses molecular signatures known or predicted to have phenotypic implications and worldwide epidemiological relevance (WHO, 2021). By July 2021, VOC Delta was the most represented lineage in the SARS-CoV-2 global pandemic profile (GISAID, 2021). VOC Delta also exhibits multiple molecular signatures in its Spike protein (T19R, E156G, Δ157-158, L452R, T478K, P681R, and D950N) (ECDC, 2021), with immunological (Q. Li et al., 2020), clinical and epidemiological implications (Johnson et al., 2020; Arora et al., 2021).

Brazil registered its first COVID-19 case in late February 2020 (Ministério da Saúde, 2021a). Since then, the country has been heavily affected by the pandemic, with more than 20 million cases and 600 thousand confirmed deaths by early October 2021 (Ministério da Saúde, 2021b). From February until June 2021, the epidemic profile across the country has been largely dominated by the VOC Gamma (P.1+P.1.*) (Rede Genômica Fiocruz 2021, http://www.genomahcov.fiocruz.br/dashboard/) that emerged in the Northern Brazilian state of Amazonas in late November 2020 (Naveca et al., 2021; Faria et al., 2021). Since June 2021, however, the VOC Delta has been occupying increasingly more significant shares of the country’s epidemic profile. By early September 2021, most Brazilian sampled genomes were assigned to the B.1.617.2 or AY.* PANGO lineages in the country’s Southeastern (87%), Southern (73%), and Northeastern (52%) regions, while VOC Gamma still dominates in the Northern (86%), and Central-Western (79%) regions (Rede Genômica Fiocruz, 2021).

The first imported case of the VOC Delta in Brazil was identified in the Southern region by late April 2021 in an event epidemiologically linked to a Brazilian with travel history through Asia in previous weeks (Agência Brasil, 2021). This first case was one of the many independent VOC Delta introductions to be detected in the country during the following months, some already producing, by April 2021, evidence of autochthonous transmission (Patané et al., 2021). In July 2021, the first large local cluster of the Delta variant was detected in the Southeastern state of Rio de Janeiro (Lamarca et al., 2021). Previous analyses of Delta in Brazil primarily focus on the most populated states of Rio de Janeiro and São Paulo. In contrast, information about the dissemination dynamics of the Delta variant in other states is scarce. In this work, we aimed to understand the emergence and spread of the VOC Delta in Brazil by inferring the variant’s phylogenetic profile across the country’s five geographical regions and determining its major dissemination routes.

## METHODOLOGY

### Dataset Composition

A multicentric effort from the COVID-19 Fiocruz Genomic Surveillance network in Brazil (http://www.genomahcov.fiocruz.br/) recovered VOC Delta complete genomes (n = 482) across 20 of 26 Brazilian states. The majority of the SARS-CoV-2 VOC Delta (B.1.617.2 + AY.*) genomes were newly generated using the Illumina COVIDSeq Test kit with the adaptation of some primers to minimize regions with drop-out (Naveca et al., 2021) or previously published in-house sequencing protocols (Resende et al., 2021, Nascimento et al., 2020). Obtained FASTQ reads were imported into the CLC Genomics Workbench v.20.0.4 (Qiagen A/S, Denmark), trimmed, and mapped against a reference sequence (EPI_ISL_402124) available in EpiCoV database of the GISAID initiative (https://www.gisaid.org/) (Elbe & Buckland-Merrett, 2017) or the consensus sequences were generated by the Viralflow (Dezordi et al., 2021). All genomes were uploaded to the EpiCoV database in GISAID (Appendix 2). This study was approved by the Ethics Committee of the Amazonas State University (CAAE: 25430719.6.0000.5016) and by the Ethics Committee of the FIOCRUZ (CAAE: 68118417.6.0000.5248).

Additionally, we downloaded all VOC Delta (B.1.617.2 + AY.*) complete genomes available at the EpiCoV database in GISAID (Elbe & Buckland-Merrett, 2017) until 2021-09-07 (n = 948,925). Sequences without a complete collection date were removed, as well as those above a threshold of unidentified positions (Ns > 3% for Brazilian sequences, and Ns > 1% for those outside the country). The retained dataset had its dimension significantly scaled-down firstly by removing identical sequences by their clustering with CD-HIT v.4.8.1 (Li et al., 2001), and subsequently by a local BLAST (Altschul et al., 1990) search against Brazilian sequences. The resulting dataset (n = 4,260) was aligned with MAFFT v7.453 (Katoh, 2002) and assigned to PANGO lineages (Rambaut et al., 2020) by the Pangolin algorithm (O’Toole et al., 2021).

### Identification of Phylogenetic Clusters

An initial dataset of 4,260 sequences was employed in a Maximum Likelihood (ML) phylogenetic reconstruction with IQ-TREE v2.1.3 (Minh et al., 2020), using a GTR + I + F + Г4 nucleotide substitution model as selected by ModelFinder (Kalyaanamoorthy et al., 2017) to identify possible phylogenetic clusters of VOC Delta in Brazilian territory. The reliability of the obtained tree topology was estimated with the approximate likelihood-ratio test (Anisimova & Gascuel, 2006). Two rounds of ML analysis were performed to improve the presentation of the results and the resolution of the inferred tree. After the first one, all statistically supported phylogenetic clusters (aLRT ≥ 75) without Brazilian sequences were removed. Brazilian phylogenetic clusters in the ML tree were defined as any statistically supported (aLRT ≥ 75) group of more than one sequence mostly (≥ 75%) originated in one of the country’s five regions.

### Phylogeographic Analysis

Two major Delta transmission clusters detected in the Southern and southeastern regions were selected for further analysis alongside a few of their basal non-Brazilian sequences. To trace back their MRCA (Most Recent Common Ancestor) and reconstruct their spatial diffusion pattern, a time-scaled phylogenetic tree was inferred in the Bayesian Markov Chain Monte Carlo (MCMC) approach as implemented in the software package BEAST v1.10.4 (Drummond et al., 2002, Marc A Suchard et al., 2018) with BEAGLE (Suchard & Rambaut, 2009) to improve run-time efficiency. Bayesian MCMC analyses were performed using a strict molecular clock model, a constant prior distribution on the substitution rate (8–10 × 10-4 substitutions per site per year), and the nonparametric Bayesian skyline model as a coalescent tree prior (Drummond et al., 2005). Migration events were reconstructed using a reversible discrete phylogeographic model (Lemey et al., 2009) with a CTMC rate reference prior (Ferreira & Suchard, 2008). MCMC chains were run for 200 × 106 generations, and convergence and uncertainty of parameter estimates were assessed by calculating the Effective Sample Size (ESS) and 95% Highest Probability Density (HPD) values with Tracer v1.7.1 (Rambaut et al., 2018). The maximum clade credibility (MCC) trees were summarized with TreeAnnotator v1.10.4 and visualized with FigTree v1.4.4 (Rambaut, 2009).

### Molecular Signatures

The Nextclade (*https://github.com/nextstrain/nextclade*) algorithm was used to access the molecular composition of Brazilian genomes. All non-synonymous substitutions and deletions detected in the majority of sequences (≥ 75%) were represented in a heatmap generated with the ggplot2 package (https://ggplot2.tidyverse.org/reference/ggplot.html).

## RESULTS

The ML phylogenetic analysis of 2,264 Delta sequences from Brazil and a selected subset of 591 non-Brazilian sequences revealed a large number (≥ 60) of discernible independent introductions in Brazil, mainly in the Southeastern and Southern regions (**Figure 1A**). Most introductions resulted in clusters of small size (2 ≤ n <20) that were composed almost exclusively of Brazilian sequences (≥ 99%), including the Brazilian cluster (*n* = 5) associated with the Delta index case in the state of Paraná (PR-I) (**Fig 1A and Table 1**). Three founder events, by contrast, resulted in major Brazilian clusters designated as BR-I (n = 1,560), BR-II (n = 208), and BR-III (n = 171) **(Fig 1A)**. According to pangolin v3.1.16, BR-I and BR-II were composed of B.1.617.2 variant sequences, and BR-III, of AY.46.3. Sampled genomes from Brazil’s Southeastern region make up the majority (≥ 90%) of the clusters BR-I and BR-III. In comparison, those from the Southern region represent the largest part (≥ 75%) of cluster BR-II. Clusters BR-I and BR-II were detected at different Brazilian states, while BR-III was mainly restricted to the São Paulo state **(Table 1)**. BR-I was the most represented cluster among genomes from Brazil’s Northern (91%), Southeastern (73%), Northeastern (71%), and Central-Western (60%) regions, and BR-II was the most prevalent cluster in the Southern region (69%) **(Fig 1B and Table S1)**.

**TABLE 1:**
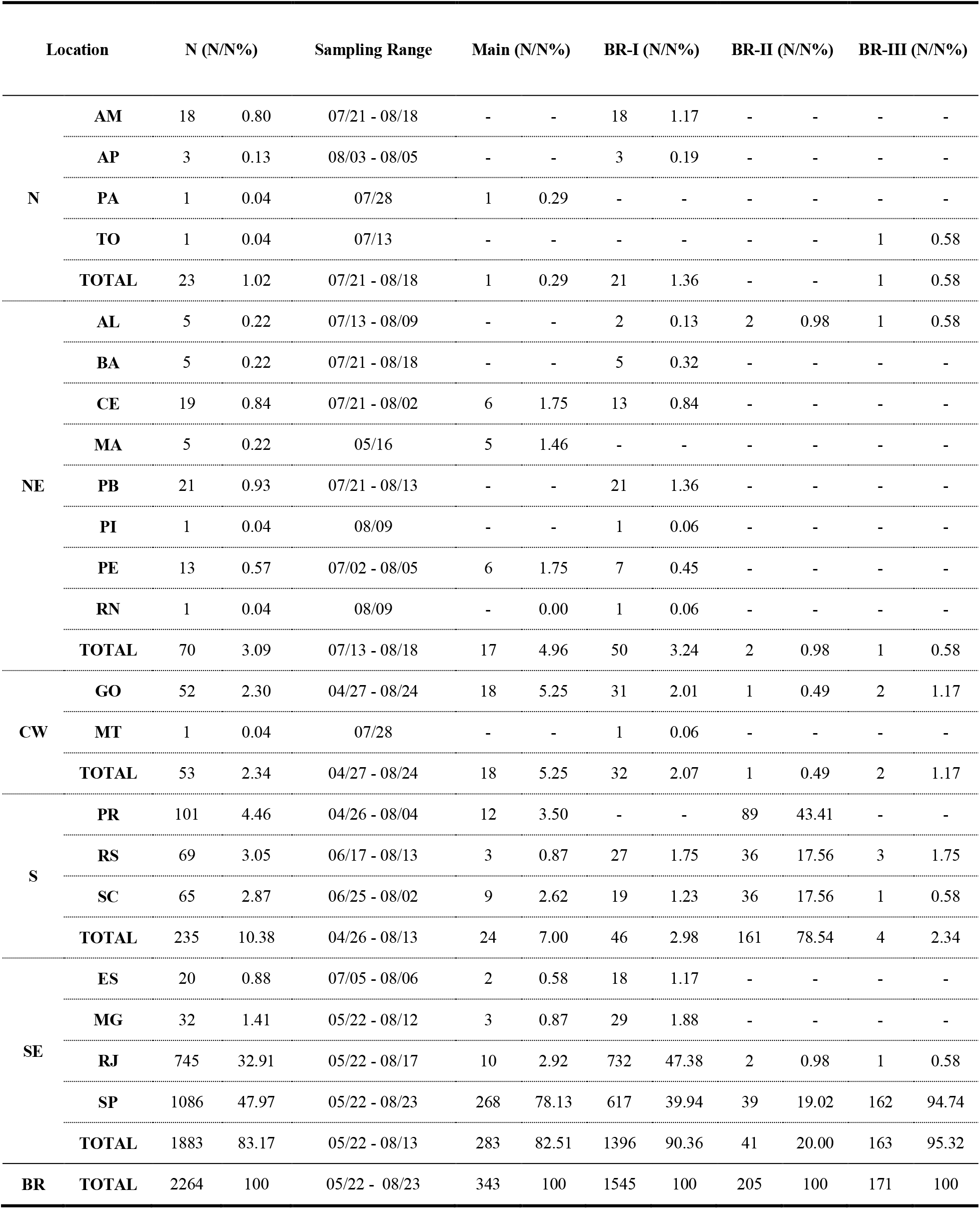
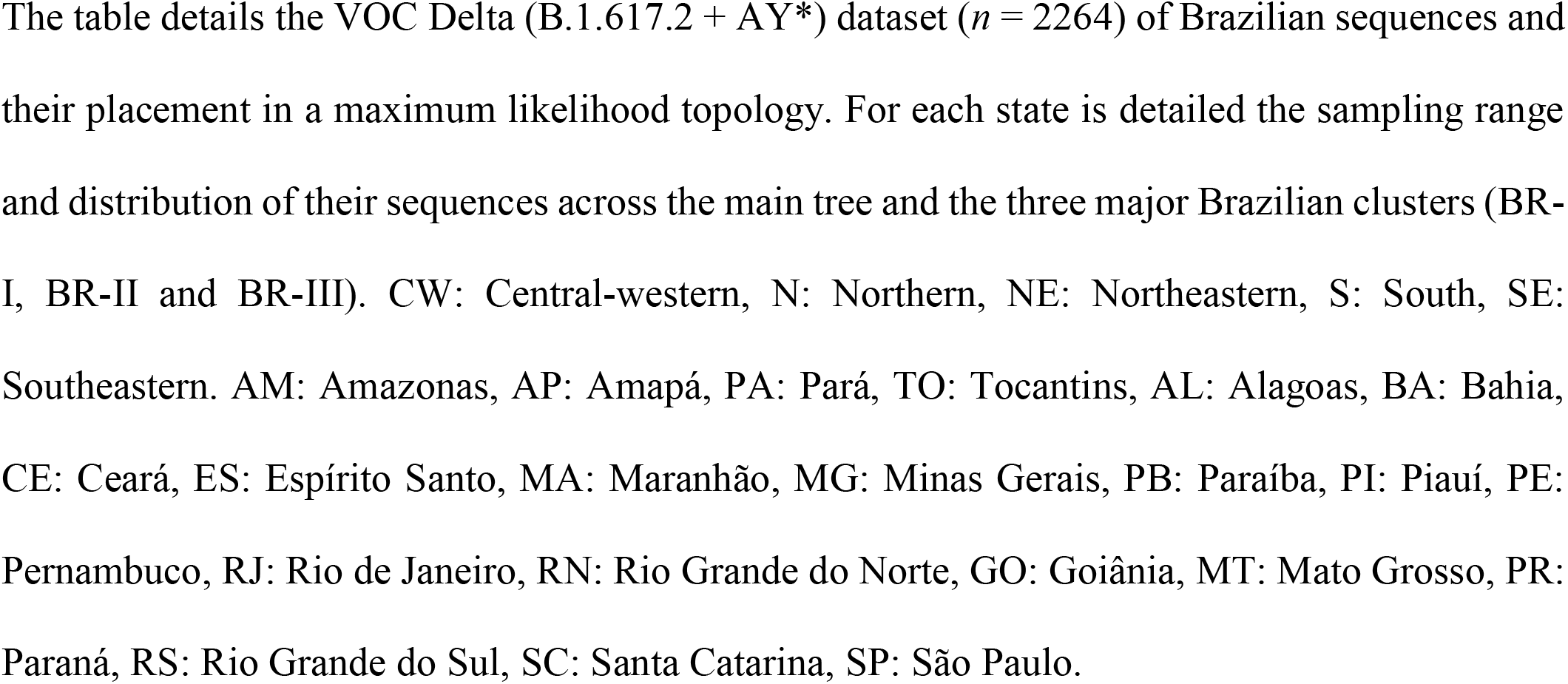
SARS-CoV-2 VOC Delta Brazilian Dataset.

| Location |  | N (N/N%) |  | Sampling Range | Main (N/N%) |  | BR-I (N/N%) |  | BR-II (N/N%) |  | BR-III (N/N%) |  |
| --- | --- | --- | --- | --- | --- | --- | --- | --- | --- | --- | --- | --- |
| N | AM | 18 | 0.80 | 07/21 - 08/18 | - | - | 18 | 1.17 | - | - | - | - |
|  | AP | 3 | 0.13 | 08/03 - 08/05 | - | - | 3 | 0.19 | - | - | - | - |
|  | PA | 1 | 0.04 | 07/28 | 1 | 0.29 | - | - | - | - | - | - |
|  | TO | 1 | 0.04 | 07/13 | - | - | - | - | - | - | 1 | 0.58 |
|  | TOTAL | 23 | 1.02 | 07/21 - 08/18 | 1 | 0.29 | 21 | 1.36 | - | - | 1 | 0.58 |
| NE | AL | 5 | 0.22 | 07/13 - 08/09 | - | - | 2 | 0.13 | 2 | 0.98 | 1 | 0.58 |
|  | BA | 5 | 0.22 | 07/21 - 08/18 | - | - | 5 | 0.32 | - | - | - | - |
|  | CE | 19 | 0.84 | 07/21 - 08/02 | 6 | 1.75 | 13 | 0.84 | - | - | - | - |
|  | MA | 5 | 0.22 | 05/16 | 5 | 1.46 | - | - | - | - | - | - |
|  | PB | 21 | 0.93 | 07/21 - 08/13 | - | - | 21 | 1.36 | - | - | - | - |
|  | PI | 1 | 0.04 | 08/09 | - | - | 1 | 0.06 | - | - | - | - |
|  | PE | 13 | 0.57 | 07/02 - 08/05 | 6 | 1.75 | 7 | 0.45 | - | - | - | - |
|  | RN | 1 | 0.04 | 08/09 | - | 0.00 | 1 | 0.06 | - | - | - | - |
|  | TOTAL | 70 | 3.09 | 07/13 - 08/18 | 17 | 4.96 | 50 | 3.24 | 2 | 0.98 | 1 | 0.58 |
| CW | GO | 52 | 2.30 | 04/27 - 08/24 | 18 | 5.25 | 31 | 2.01 | 1 | 0.49 | 2 | 1.17 |
|  | MT | 1 | 0.04 | 07/28 | - | - | 1 | 0.06 | - | - | - | - |
|  | TOTAL | 53 | 2.34 | 04/27 - 08/24 | 18 | 5.25 | 32 | 2.07 | 1 | 0.49 | 2 | 1.17 |
| S | PR | 101 | 4.46 | 04/26 - 08/04 | 12 | 3.50 | - | - | 89 | 43.41 | - | - |
|  | RS | 69 | 3.05 | 06/17 - 08/13 | 3 | 0.87 | 27 | 1.75 | 36 | 17.56 | 3 | 1.75 |
|  | SC | 65 | 2.87 | 06/25 - 08/02 | 9 | 2.62 | 19 | 1.23 | 36 | 17.56 | 1 | 0.58 |
|  | TOTAL | 235 | 10.38 | 04/26 - 08/13 | 24 | 7.00 | 46 | 2.98 | 161 | 78.54 | 4 | 2.34 |
| SE | ES | 20 | 0.88 | 07/05 - 08/06 | 2 | 0.58 | 18 | 1.17 | - | - | - | - |
|  | MG | 32 | 1.41 | 05/22 - 08/12 | 3 | 0.87 | 29 | 1.88 | - | - | - | - |
|  | RJ | 745 | 32.91 | 05/22 - 08/17 | 10 | 2.92 | 732 | 47.38 | 2 | 0.98 | 1 | 0.58 |
|  | SP | 1086 | 47.97 | 05/22 - 08/23 | 268 | 78.13 | 617 | 39.94 | 39 | 19.02 | 162 | 94.74 |
|  | TOTAL | 1883 | 83.17 | 05/22 - 08/13 | 283 | 82.51 | 1396 | 90.36 | 41 | 20.00 | 163 | 95.32 |
| BR | TOTAL | 2264 | 100 | 05/22 - 08/23 | 343 | 100 | 1545 | 100 | 205 | 100 | 171 | 100 |

The table details the VOC Delta (B.1.617.2 + AY\*) dataset ( $n = 2264$ ) of Brazilian sequences and their placement in a maximum likelihood topology. For each state is detailed the sampling range and distribution of their sequences across the main tree and the three major Brazilian clusters (BR-I, BR-II and BR-III). CW: Central-western, N: Northern, NE: Northeastern, S: South, SE: Southeastern. AM: Amazonas, AP: Amapá, PA: Pará, TO: Tocantins, AL: Alagoas, BA: Bahia, CE: Ceará, ES: Espírito Santo, MA: Maranhão, MG: Minas Gerais, PB: Paraíba, PI: Piauí, PE: Pernambuco, RJ: Rio de Janeiro, RN: Rio Grande do Norte, GO: Goiânia, MT: Mato Grosso, PR: Paraná, RS: Rio Grande do Sul, SC: Santa Catarina, SP: São Paulo.

**FIGURE 1.**
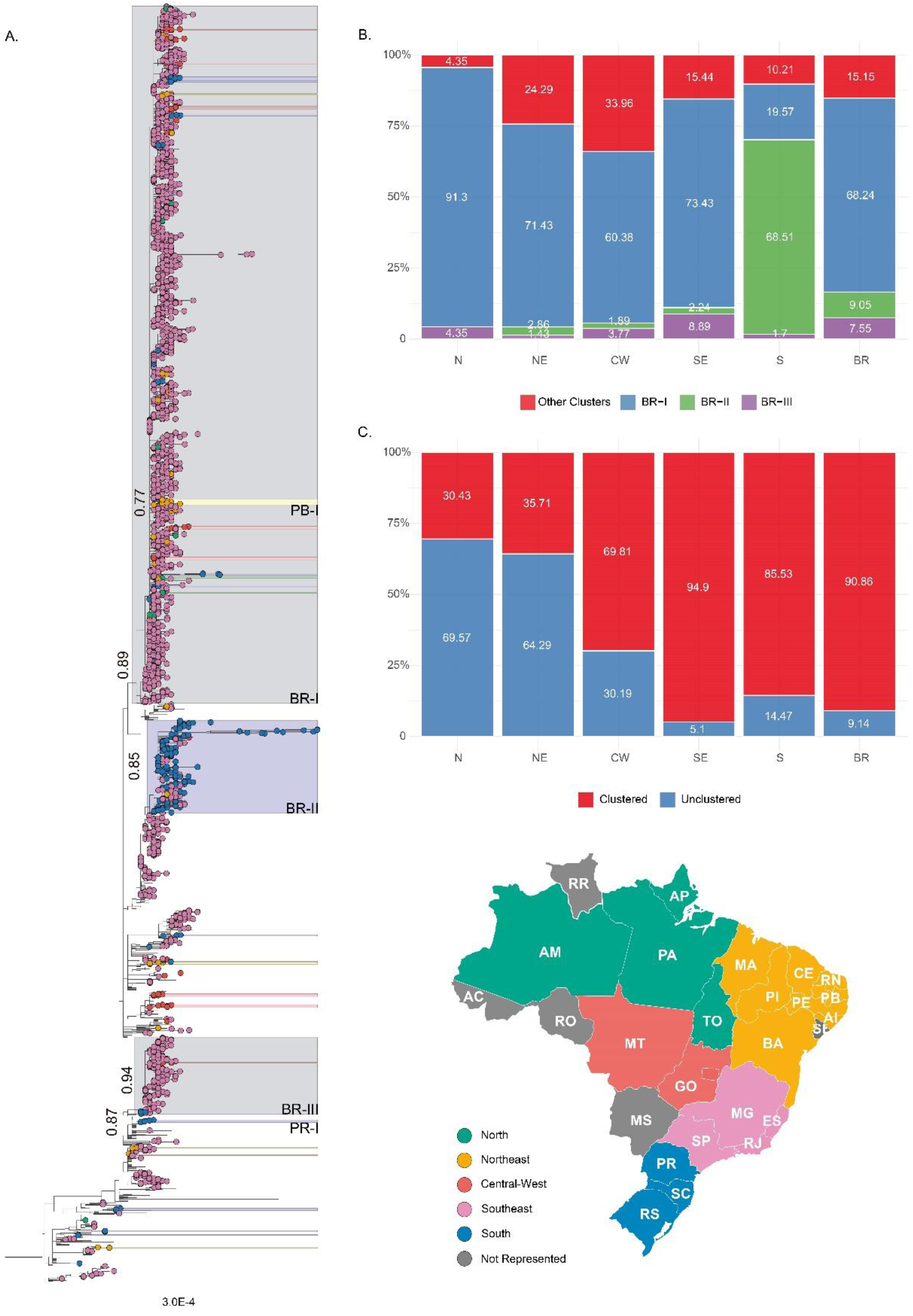
Emergence of SARS-CoV-2 VOC Delta Brazilian Phylogenetic Clusters. A. Maximum Likelihood (ML) tree constructed with Brazilian SARS-Cov-2 VOC Delta (B.1.617.2 + AY*) genomes (n = 2,855). Tip shapes are colored according to the map in the bottom left of their Brazilian geographic region of origin. States not represented in the dataset are colored in dark grey. All statistically supported (aLRT ≥ 0.75) groups independent of their dimension have the same scholar scheme. Two of Brazilians’ main clusters (BR-I and BR-II), composed of southeastern sequences, are colored in light gray. Sequences outside Brazil had their tips removed for improved clarity. The three main clusters (BR-I, BR-II, and BR-III) and other noticeable clusters are named, and their statistical support (aLRT) is also indicated. B. Distribution of sampled genomes from Brazil (BR, n = 2,264) and its North (N, n = 23), Northeast (NE, n = 70), Central-western (CW, n = 53), Southeast (SE, n = 1,883), and South (S, n = 235) regions across the three main clusters (BR-I, BR-II, and BR-III) and the main inferred ML VOC Delta tree (Main). B. Distribution of sampled genomes from Brazil and its five regions across the ML tree clusterization profile in unclustered and clustered, a category defined by any statistical supported (aLRT ≥ 0.75) grouping of more than one sequence from the same geographical region.

Most Delta sequences sampled in the Southeastern (95%) and Southern (86%) regions branched within Brazilian clusters, while much lower levels of clusterization were detected in the other country regions (**Figure 1C**). The Northern Brazilian region was the least represented in the national Delta dataset (n = 23, 1%), harbored samples from four of the six region’s states, and had the lowest level of local clusterization (30%). Amazonas was the most represented state (n = 18, 0.8%) **(Table 1)**, with samples collected since late July 2021, and displayed the largest Delta cluster in the region (n = 3 sequences). The Northeastern region (n = 70, 3%) harbored samples from eight of the nine region’s states, the most represented ones being Paraíba (n = 21), Ceará (n = 19) and Pernambuco (n = 13) **(Table 1)**. Despite a larger number of sequences than Brazil’s Northern region, the clusterization level observed (36%) was similar in both regions. The largest Delta cluster in the Northeastern region was detected in the Paraíba state (PB-I, n = 12 sequences) and comprises 57% of Delta sequences from that state **(Fig 1A)**. The Central-West region (n = 53, 2%) comprehends sequences from two of three states, being nearly all recovered from the Goiás state since late July (n = 52) **(Table 1)**. The observed level of clusterization in Goiás state (70%) was significantly higher than in Northern and Northeastern regions. However, Delta sequences were highly scattered in the tree’s topology, forming multiple (n = 11) clusters of small size (2 ≤ n ≤7 sequences).

While the dissemination dynamics of Delta cluster BR-I was already explored in previous studies (Lamarca et al., 2021; Patané et al., 2021), the spatiotemporal pattern of clusters BR-II and BR-III remained unknown. Bayesian phylogeographic analysis supports that Paraná was the most probable source location of cluster BR-II [*PSP* = 0.57] at late April 2021 (median = 2021-04-27, 95% HPD: 2021-03-21/2021-05-17) (**Fig 2A**). Paraná was the main dissemination hub of cluster BR-II, followed by Santa Catarina. Most BR-II genomes hosted the molecular synapomorphies ORF3a:A23V (100%), ORF1a:A3070V (98%) and S:T95I (85%) (**Fig 2C**). The MRCA of cluster BR-III was traced back to early June 2021 (median = 2021-06-04, 95% HPD: 2021-05-13/2021-06-20) in the São Paulo state [PSP = 0.58] **(Fig 2B)**. São Paulo was the main hub of dissemination of cluster BR-III. Despite being detected in seven states, the dissemination of cluster BR-III outside São Paulo was highly limited and mostly reduced to single introductions, except for one small cluster in Goiás (n = 2) [PSP = 1.0]. The BR-III genomes had one molecular synapomorphy, ORF9b:R32L **(Fig 2C)**.

**FIGURE 02.**
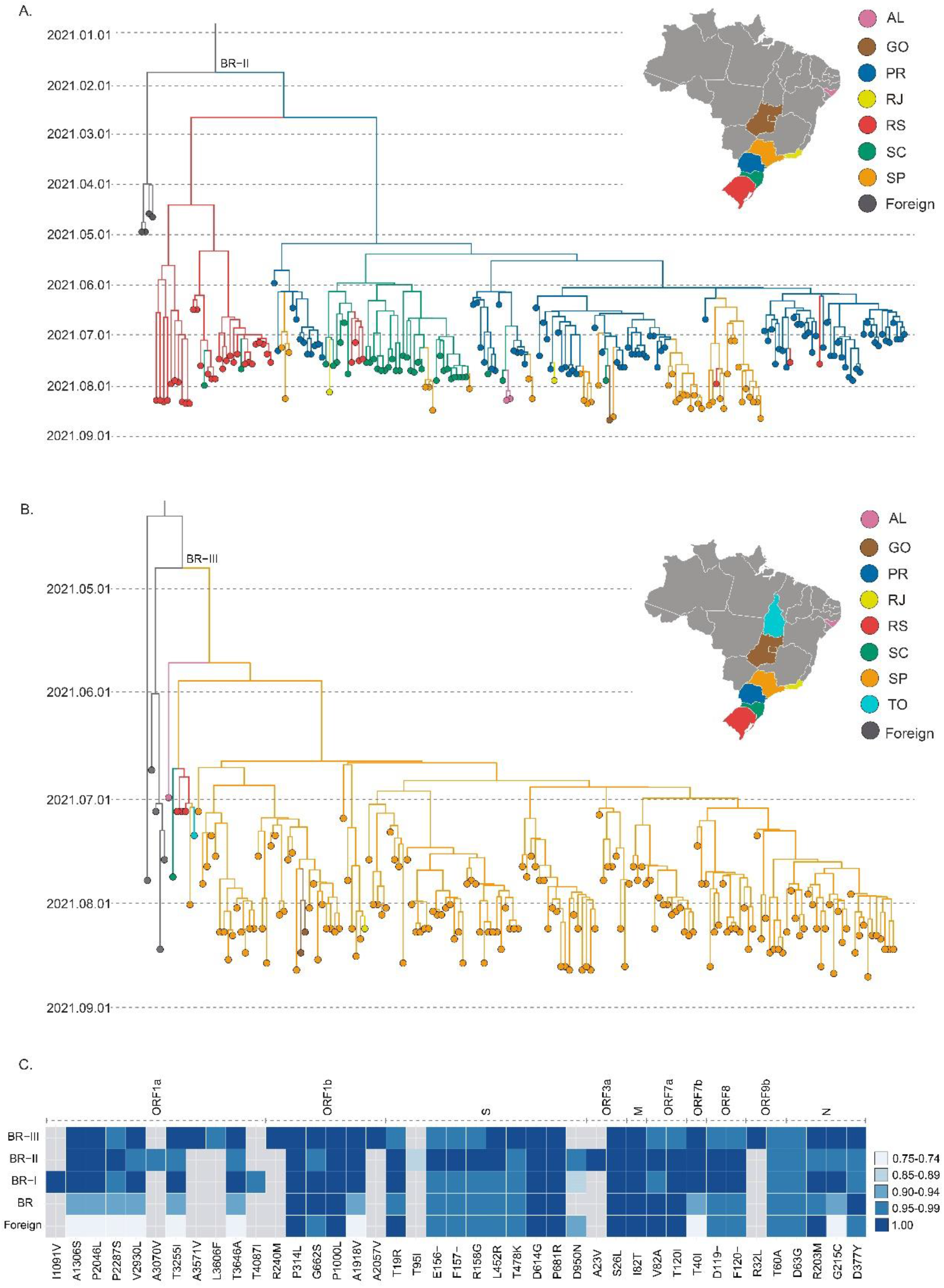
Spatial, Temporal and Molecular Characterization of VOC Delta BR-II and BR-III Clusters. A-B. Time-scaled MCC tree of SARS-Cov-2 VOC Delta (B.1.617.2 + AY*) BR-II cluster (*n* = 207) (A) and BR-III (*n* = 171) (B), one of the main Brazilian phylogenetic clusters of this variant. Branches are colored according to their most probable location, based on the color scheme shown in the upper right corner legend. C. Description of molecular signatures of the BR-II Cluster. Lines indicated by BR-I (*n* = 1,560), BR-II (*n* = 208), and BR-III (*n* = 171) summarize, in comparison to the Wuhan 2019 reference sequence, the relative frequency of non-synonymous substitutions and deletions observed in the majority (≥ 75%) of sequences composing each cluster. The BR line represents the same procedure applied to sequences from Brazil outside the three main clusters (*n* = 325), and the foreign line to the ones outside Brasil is used as referenced in the complete ML tree (*n* = 591). The substitution and its gene are annotated in the bottom and upper margins, respectively. Frequencies are represented according to the legend on the right. AL: Alagoas, GO: Goiás, PR: Paraná, RS: Rio Grande do Sul, SC: Santa Catarina, SP: São Paulo, TO: Tocantins.

## DISCUSSION

In this work, we explored the emergence and spread of the VOC Delta in different Brazilian regions. The cluster BR-I, previously associated with the large local cluster of the Delta variant in the Southeastern state of Rio de Janeiro (Lamarca et al., 2021), was here confirmed as the main driver behind the variant expansion in several Brazilian states of the Northern, Northeastern and Central-Western regions. The Delta epidemic in the Southern region, however, was mainly driven by the cluster BR-II. A third transmission cluster of large size here named BR-III was also identified mainly in São Paulo.

The Southeastern Brazilian region contributed with 83% (n = 1,833) of the VOC Delta sequences here analyzed, followed by Southern region with 10% (n = 235), Northeastern region with 3% (n = 70), Central-Western region with 2% (n = 53) and Northern region with 1% (n = 23). This distribution mirrors the relative prevalence of VOC Delta across regions and also closely resembles the inferred clusterization levels. The proportion of sequences in each region with statistically supported association to another sampled genome from the same region was used as a proxy of local transmission events in opposition to independent introductions. The Southeastern and Southern regions displayed the more sustained VOC Delta epidemics with large fractions of their sampled genomes (86% - 95%) clusterized among each region’s dataset. On the other hand, lower levels of clusterization (30-70%) were observed in the remaining regions.

Our data support that Brazilian states presents Delta epidemics in different stages of maturity. The oldest and widely established Delta epidemics were observed in the Southeastern and Southern regions. The Delta epidemic in Goiás (Central-West region), as in most of the states from the Northeastern region, is currently driven by multiple clusters of small size, characteristic of an intermediate stage. The most notable exception was the state of Paraiba that housed the largest phylogenetic cluster of Delta in the Northeastern region, and this cluster comprises more than 50% of the Delta sequences from that state. The Delta epidemic in the Northern region seems to be at the very early dissemination stage as most sequences from this region (70%) appeared unclustered with almost no evidence of local spread. Continuous genomic monitoring in these states is essential, as these are likely candidates in becoming secondary hubs of VOC Delta dissemination in their respective regions.

The cluster BR-II was the primary responsible for disseminating the VOC Delta in Brazil’s Southern region, aggregating a significant fraction of its samples (69%) and was responsible for an even more significant fraction of samples from Paraná state (88%), the most probable source location. Other sampled genomes from Paraná were mostly aggregated in small clusters (n = 3), and no introduction of the cluster BR-I was detected in the state, even though BR-I has multiple sampled genomes in the other two southern states, Rio Grande do Sul (n = 27) and Santa Catarina (n = 19). This result could indicate that BR-II dominates the VOC Delta epidemic in the Paraná state, limiting the spread of other clusters in the same network of susceptible individuals. Given the date the MRCA of cluster BR-II was traced back to late April 2021, close to the first detection of the VOC Delta in the country, a founder event effect could be associated with the successful dissemination of this cluster across Brazil’s south region.

The cluster BR-II exhibits ORF3a:A23V as a molecular synapomorphy. The ORF3a gene is an ion transporter of 275 amino acids (Ren et al., 2020), that when up regulated, leads to an increased fibrinogen secretion associated with the COVID-19 characteristic cytokine storm (Tan et al., 2005). The ORF3a activates the NLRP3 inflammasome by promoting TRAF3 (TNF receptor-associated factor 3)–mediated ubiquitination of apoptosis-associated speck-like protein with a caspase recruitment domain (Siu et al., 2019). The position identified here is located near the ORF3A:TRAF3 interaction site (Timmers et al., 2021), raising the possibility of eventual phenotypic implications. Among the three sites (ORF1a:A3070V, S:T95I, and ORF3a:A23V) with higher prevalence in the cluster BR-II, ORF3a:A23V presence is significantly distinct to other Brazilian major clusters and foreign reference sequences. The ORF3a:A23V mutation was a unique signature observed in all sequences within this cluster but not found in the other genomes in our dataset.

The cluster BR-III was the most recently emerged of the three main Brazilian Delta clusters, having its MRCA traced back to early June. Most genomes sampled outside São Paulo (*n* = 6) were aggregated in the base of the cluster, which could be associated with the inferred low support for BR-III MRCA location in São Paulo [*PSP* = 0.58]. Despite being heavily composed of sequences from São Paulo (≥ 90%), all Brazilian regions were represented in the BR-III dataset. The BR-III clade also exhibited one molecular synapomorphy, ORF9b:R32L. ORF9b product localizes in the membrane of the mitochondria, acting in the inhibition of IFN-1 secretion (Jiang et al., 2020).

The clusters BR-I to BR-III comprise the majority (85%) of Delta sequences here analyzed, while most other Brazilian sequences represented dead-end introductions or unsustained transmission clusters. Among these limited outbreaks was Brazil’s initially identified cluster PR-I (*n* = 5) associated with the country’s index case located in Paraná (Patané et al., 2021) and two additional limited outbreaks registered in Brazil’s Northeastern region associated with marine vessels: one in the state of Maranhão (*n* = 5 sequences) returning from Malaysia in middle May 2021 and the other in the state of Pernambuco (*n* = 5 sequences) in early June 2021 (Patané et al., 2021). The lack of evidence of further transmission of these Delta clusters supports the success of preventive measures implemented in Brazil to contain such initial introductions.

This work is certainly limited by the non-availability of more robust metadata and possibly insufficient sampling in several Brazilian states. VOC Delta is relatively susceptible to the available vaccines (Liu et al., 2021; Lustig et al., 2021). Indeed, information regarding vaccination status would allow for the elucidation of potentially conditioning factors in establishing local clusters in Brazil. In our dataset, five states were not represented by any sequence, Acre, Roraima, and Rondônia in the Northern, Mato Grosso do Sul in the Central-West, and Sergipe in the Northeast. In contrast, other states were represented by very few (n < 10) sequences. A more globally strenuous sampling could lead to the elucidation of dissemination paths and identification of new clusters across Brazilian states with lower Delta prevalence. This effort would allow for more promptness in the response of public health authorities in their control of new introductions.

In the rapid turnover of variants characteristic of the SAR-CoV-2 pandemic, Brazilian regions seem to occupy different stages of an increasingly more significant participation of the VOC Delta in their epidemic profiles. This process demands continuous genomic and epidemiologic surveillance to identify and mitigate new introductions, limit their dissemination, and prevent the establishment of massive outbreaks in a population already heavily affected by the COVID-19 pandemic.

## Data Availability

All genomes were uploaded to the EpiCoV database in GISAID (https://www.gisaid.org/).

## AUTHORS CONTRIBUTIONS

IA, FGN, TG, ED, MMS, GB, and PCR conceived and designed the study. FGN, FM, HF, GLW collected the samples and worked on sequence protocols. LRA, ECP, TMMV, ASR and RSL, worked on sequencing protocols. IA and PCR performed the phylogenetic and phylodynamics inferences. IA, GB, and PCR wrote the first draft of the manuscript. All authors reviewed and approved the final version of the manuscript.

## ACKNOWLEDGEMENTS

The authors wish to thank all the health care workers and scientists who have worked hard to deal with this pandemic threat, the GISAID team, and all the EpiCoV database’s submitters. GISAID acknowledgment table containing sequences used in this study is shown in Supplementary Table 3. We also appreciate the support of the Fiocruz COVID-19 Genomic Surveillance Network (http://www.genomahcov.fiocruz.br/) members, the Respiratory Viruses Genomic Surveillance Network of the General Laboratory Coordination (CGLab), Brazilian Ministry of Health (MoH), Brazilian Central Laboratory States (LACENs), and the Amazonas surveillance teams for the partnership in the viral surveillance in Brazil.

## FUNDING

Financial support was provided by Fundação de Amparo à Pesquisa do Estado do Amazonas (FAPEAM) (PCTI-EmergeSaúde/AM call 005/2020 and Rede Genômica de Vigilância em Saúde-REGESAM); Conselho Nacional de Desenvolvimento Científico e Tecnológico (CNPq) (grant 402457/2020–0); CNPq/Ministério da Ciência, Tecnologia, Inovações e Comunicações/Ministério da Saúde (MS/FNDCT/SCTIE/Decit) (grant 403276/2020-9); Departamento da Ciência e Tecnologia (DECIT), Ministério da Saúde; Inova Fiocruz/Fundação Oswaldo Cruz (Grants VPPCB-007-FIO-18–2–30 and VPPCB-005-FIO-20–2–87), INCT-FCx (465259/2014–6) and Fundação Carlos Chagas Filho de Amparo à Pesquisa do Estado do Rio de Janeiro (FAPERJ) (26/210.196/2020). This work was also supported by the Pan American Health Organization (PAHO), Brazil Country Office. F.G.N, G.L.W, G.B and M.M.S are supported by the CNPq through their productivity research fellowships (306146/2017–7, 303902/2019–1, 302317/2017–1 and 313403/2018-0, respectively). G.B. is also funded by FAPERJ (Grant number E-26/202.896/2018).

## Notes

### Competing Interest Statement

The authors have declared no competing interest.

### Funding Statement

Financial support was provided by Fundacao de Amparo a Pesquisa do Estado do Amazonas (FAPEAM) (PCTI EmergeSaude/AM call 005/2020 and Rede Genomica de Vigilancia em Saude REGESAM); Conselho Nacional de Desenvolvimento Cientifico e Tecnologico (CNPq) (grant 402457/2020 0); CNPq/Ministerio da Ciencia, Tecnologia, Inovacoes e Comunicacoes/Ministerio da Saude (MS/FNDCT/SCTIE/Decit) (grant 403276/2020 9); Departamento da Ciencia e Tecnologia (DECIT), Ministerio da Saude; Inova Fiocruz/Fundacao Oswaldo Cruz (Grants VPPCB 007 FIO 18 2 30 and VPPCB 005 FIO 20 2 87), INCT FCx (465259/2014 6) and Fundacao Carlos Chagas Filho de Amparo a Pesquisa do Estado do Rio de Janeiro (FAPERJ) (26/210.196/2020). This work was also supported by the Pan American Health Organization (PAHO), Brazil Country Office. F.G.N, G.L.W, G.B and M.M.S are supported by the CNPq through their productivity research fellowships (306146/2017 7, 303902/2019 1, 302317/2017 1 and 313403/2018 0, respectively). G.B. is also funded by FAPERJ (Grant number E 26/202.896/2018).

### Author Declarations

This study was approved by the Ethics Committee of the Amazonas State University (CAAE: 25430719.6.0000.5016) and by the Ethics Committee of the FIOCRUZ (CAAE: 68118417.6.0000.5248).

