## Supplementary Material for "Emergence and Spread of the SARS-CoV-2 Variant of Concern Delta Across Different Brazilian Regions"

**SUPPLEMENTARY TABLES**

**SUPPLEMENTARY TABLE 1: Distribution SARS-CoV-2 VOC Delta Dataset from Brazil's Five regions Across the Country's Three Main Clusters**

| Cluster | BR (N/N%) | | N (N/N%) | | NE (N/N%) | | CW (N/N%) | | SE (N/N%) | | S (N/N%) | |
|---|---|---|---|---|---|---|---|---|---|---|---|---|
| **Main** | 343 | 15,15 | 1 | 4.35 | 17 | 24.29 | 18 | 33.96 | 283 | 15.44 | 24 | 10.21 |
| **BR-I** | 1545 | 68,24 | 21 | 91.30 | 50 | 71.43 | 32 | 60.38 | 1,396 | 73.43 | 46 | 19.57 |
| **BR-II** | 205 | 9,05 | 0 | 0.00 | 2 | 2.86 | 1 | 1.89 | 41 | 2.24 | 161 | 68.51 |
| **BR-III** | 171 | 7,55 | 1 | 4.35 | 1 | 1.43 | 2 | 3.77 | 163 | 8.89 | 4 | 1.70 |
| **Total** | 2264 | 100 | 23 | 100 | 70 | 100 | 53 | 100 | 1,883 | 100 | 235 | 100 |

The table details the distribution of sampled genomes from Brazil's Five regions across its SARS-CoV-2 VOC Delta main three and major clusters, BR-I (*n* = 1,560), BR-II (*n* = 207), and BR-III (*n* = 171). BR: Brazil, CW: Central-western, N: North, NE: Northeast, SE: Southeast, S: South.

**SUPPLEMENTARY TABLE 2: SARS-CoV-2 VOC Delta Clusterization Profile**

| Clusterization | BR (N/N%) | | N (N/N%) | | NE (N/N%) | | CW (N/N%) | | SE (N/N%) | | S (N/N%) | |
|---|---|---|---|---|---|---|---|---|---|---|---|---|
| **Unclustered** | 207 | 9.14 | 16 | 69.57 | 45 | 64.29 | 16 | 30.19 | 96 | 5.10 | 34 | 14.47 |
| **Clustered** | 2057 | 90.86 | 7 | 30.43 | 25 | 35.71 | 37 | 69.81 | 1787 | 94.90 | 201 | 85.53 |
| **Total** | 2264 | 100 | 23 | 100 | 70 | 100 | 53 | 100 | 1883 | 100 | 235 | 100 |

The table details the clusterization profile of genomes sampled in Brazil (BR, *n* = 2,264) and its North (N, *n* = 23), Northeast (NE, *n* = 70), Central-western (CW, *n* = 53), Southeast (SE, *n* = 1,883), and South (S, *n* = 235) regions. Phylogenetic clusters were defined as any statistically supported (aLRT ≥ 75) group of more than one sequence from the same geographical region. BR: Brazil, CW: Central-western, N: North, NE: Northeast, SE: Southeast, S: South.

### APPENDIX 1

#### COVID-19 Fiocruz Genomic Surveillance Network

| | |
|---|---|
| Carlos Leonardo Araújo<br>Cleber Furtado Akesenen<br>Fernando Braga Stehling Dias<br>Igor Oliveira Duarte<br>Jamille Mendes Bezerra<br>Joaquim Cesar Sousa Jr<br>Pedro Miguel Carneiro Jerônimo<br>Suzana Almeida Porto<br>Thaís de Oliveira Costa<br>Thais Ferreira de Oliveira<br>Ticiane Cavalcante de Souza<br>Veridiana Pessoa Miyajima | *Analytical Competence Molecular Epidemiology Laboratory (ACME), Fundação Oswaldo Cruz (FIOCRUZ) – Ceará, Brasil* |
| Acacia Lourenço Francisco Nasr<br>Ana Carolina De la Vechia<br>Rosana Aparecida Piler<br>Tatiane Motta Huggler | *Divisão de Vigilância de Doenças Transmissíveis, Secretaria Estadual de Saúde do Paraná – Paraná, Brasil* |
| Cristiano Fernandes | *Fundação de Vigilância em Saúde do Amazonas - Dra. Rosemary Costa Pinto – Amazonas, Brasil* |
| Marcelo Gomes | *Grupo de Métodos Analíticos em Vigilância Epidemiológica, Programa de Computação Científica (PROCC), Fundação Oswaldo Cruz (FIOCRUZ) – Rio de Janeiro, Brasil* |
| Adriano Abbud<br>Katia Oliveira Correa | *Instituto Adolfo Lutz – São Paulo, Brasil* |
| Alexandre Freitas da Silva<br>Antonio Marinho da Silva Neto<br>Cássia Docena<br>Filipe Zimmer Dezordi<br>Gustavo Barbosa de Lima<br>Laís Ceschini Machado<br>Lilian Caroliny Amorim Silva<br>Marcelo Henrique Santos Paiva<br>Matheus Filgueira Bezerra<br>Raul Emídio de Lima | *Instituto Aggeu Magalhães, Fundação Oswaldo Cruz (FIOCRUZ) – Pernambuco, Brasil* |
| Andreia Akemi Suzukawa<br>Mauro de Medeiros Oliveira<br>Michelle Orane Schemberger | *Instituto Carlos Chagas (ICC), Fundação Oswaldo Cruz (FIOCRUZ) – Paraná, Brasil* |
| Beatriz Grinsztejn<br>Patricia Brasil<br>Valdiléa G Veloso | *Instituto Nacional de Infectologia Evandro Chagas (INI), Fundação Oswaldo Cruz (FIOCRUZ) – Rio de Janeiro, Brasil* |
| Felicidade Pereira | *Laboratorio Central de Saúde Pública da Bahia (LACEN-BA) – Bahia, Brasil* |
| Dalane Loudal Florentino Teixeira<br>Haline Barroso | *Laboratório Central de Saúde Pública da Paraíba (LACEN-PB) – Paraíba, Brasil* |
| Anderson Brandao Leite | *Laboratório Central de Saúde Pública de Alagoas (LACEN-AL) – Alagoas, Brasil* |
| Vinicius Lemes da Silva | *Laboratorio Central de Saúde Pública de Goiás (LACEN-GO) – Goiás, Brasil* |
| André Felipe Leal Bernardes<br>Felipe Campos de Melo Iani | *Laboratório Central de Saúde Pública de Minas Gerais (LACEN-MG) – Minas Gerais, Brasil* |
| Irina Riediger<br>Maria do Carmo Debur | *Laboratorio Central de Saúde Pública de Paraná (LACEN-PR) – Paraná, Brasil* |
| | *Laboratorio Central de Saúde Pública de Pernambuco (LACEN-PE) – Pernambuco, Brasil* |
| Themis Rocha | *Laboratorio Central de Saúde Pública de Sergipe (LACEN-RN) – Rio Grande do Norte, Brasil* |
| Andreia Santos Costa<br>Lindomar dos Anjos Silva | *Laboratório Central de Saúde Pública do Amapá (LACEN-AP) – Amapá, Brasil* |
| Tirza Peixoto Mattos | *Laboratório Central de Saúde Pública do Amazonas (LACEN-AM) – Amazonas, Brasil* |
| Ana Barjud Marques Maximo<br>Liana Perdigão Mello<br>Vania Angelica Feitosa Viana | *Laboratório Central de Saúde Pública do Ceará (LACEN-CE) – Ceará, Brasil* |

| | |
|---|---|
| Rodrigo Ribeiro Rodrigues | *Laboratório Central de Saúde Pública do Espirito Santo (LACEN-ES) – Espírito Santo, Brasil* |
| Darcita Buerger Rovaris<br>Sandra Bianchini Fernandes | *Laboratorio Central de Saúde Pública do Estado de Santa Catarina (LACEN-SC) – Santa Catarina, Brasil* |
| Lidio Gonçalves Lima Neto | *Laboratório Central de Saúde Pública do Maranhão (LACEN-MA) – Maranhão, Brasil* |
| Valnete Andrade | *Laboratório Central de Saúde Pública do Pará (LACEN-PA) – Pará, Brasil* |
| Andrea Cony Cavalcanti | *Laboratório Central de Saúde Pública do Rio de Janeiro (LACEN-RJ) – Rio de Janeiro, Brasil* |
| Richard Steiner Salvato<br>Tatiana Schäffer Gregianini | *Laboratório Central de Saúde Pública do Rio Grande do Sul (LACEN-RS) – Rio Grande do Sul, Brasil* |
| Jucimária Dantas Galvão | *Laboratorio Central de Saúde Pública do Tocantins (LACEN-TO) – Tocantins, Brasil* |
| Ágatha Costa<br>André de Lima Guerra Corado<br>Fernanda Nascimento<br>George Silva<br>Karina Pessoa<br>Luciana Fé Gonçalves<br>Maria Júlia Brandão<br>Matilde Mejía<br>Michele Silva de Jesus<br>Valdinete Alves Nascimento<br>Victor Souza | *Laboratório de Ecologia de Doenças Transmissíveis na Amazônia (EDTA), Instituto Leônidas e Maria Deane, Fundação Oswaldo Cruz (FIOCRUZ) – Amazonas, Brasil* |
| Bruna Mendonça da Silva<br>Fernando do Couto Motta<br>Jéssica de Macedo Carvalho<br>Larissa Macedo Pinto | *Laboratório de Vírus Respiratórios e Sarampo, Instituto Oswaldo Cruz (IOC), Fundação Oswaldo Cruz (FIOCRUZ) – Rio de Janeiro, Brasil* |
| Fernando Vinhal | *Laboratório HLAGYN – Goiás, Brasil* |
| Isabela de Lucena Heráclio<br>Morgana de Freitas Caraciolo<br>Roberta Mendes Abreu Silva<br>Silvio Rodrigues de Almeida<br>Thayna Karoline Sousa Silva | *Programa de Treinamento em Epidemiologia Aplicada (EpiSUS-Avançado), Ministério da Saúde – Brasil* |
| Alessandro Álvares Magalhães<br>Érika Lopes Rocha Batista | *Secretaria de Saúde de Aparecida de Goiânia – Goiás, Brasil* |
| Greice Madeleine Ikeda do Carmo<br>Janaína Sallas<br>Walquiria Aparecida Almeida | *Secretaria de Vigilância em Saúde, Ministério da Saúde – Brasil* |
| Marcio Garcia<br>Cecilia Leite Costa<br>Eduardo Ruback dos Santos | *Secretaria de Vigilância em Saúde, Secretaria Municipal de Saúde – Rio de Janeiro, Brasil*<br>*Unidade de Apoio Diagnostico (UNADIG), Fundação Oswaldo Cruz (FIOCRUZ) – Ceará, Brasil* |
| João Felipe Bezerra | *Universidade Federal da Paraíba (UFPB) – Paraíba, Brasil* |

#### APPENDIX 2

#### Sequences Produced by the COVID-19 Fiocruz Genomic Surveillance Network

| | | | | | |
|---|---|---|---|---|---|
| EPI_ISL_2311861 | EPI_ISL_3536535 | EPI_ISL_3540020 | EPI_ISL_3801803 | EPI_ISL_3801843 | EPI_ISL_3801883 |
| EPI_ISL_2443545 | EPI_ISL_3539829 | EPI_ISL_3540021 | EPI_ISL_3801804 | EPI_ISL_3801844 | EPI_ISL_3801884 |
| EPI_ISL_2466268 | EPI_ISL_3539830 | EPI_ISL_3540022 | EPI_ISL_3801805 | EPI_ISL_3801845 | EPI_ISL_3801885 |
| EPI_ISL_2645411 | EPI_ISL_3539906 | EPI_ISL_3540023 | EPI_ISL_3801806 | EPI_ISL_3801846 | EPI_ISL_3801886 |
| EPI_ISL_2645412 | EPI_ISL_3539907 | EPI_ISL_3540024 | EPI_ISL_3801807 | EPI_ISL_3801847 | EPI_ISL_3801887 |
| EPI_ISL_2645413 | EPI_ISL_3539908 | EPI_ISL_3540025 | EPI_ISL_3801808 | EPI_ISL_3801848 | EPI_ISL_3801888 |
| EPI_ISL_2645414 | EPI_ISL_3539909 | EPI_ISL_3540026 | EPI_ISL_3801809 | EPI_ISL_3801849 | EPI_ISL_3801889 |
| EPI_ISL_2645415 | EPI_ISL_3539910 | EPI_ISL_3540027 | EPI_ISL_3801810 | EPI_ISL_3801850 | EPI_ISL_3801890 |
| EPI_ISL_2677318 | EPI_ISL_3539911 | EPI_ISL_3540028 | EPI_ISL_3801811 | EPI_ISL_3801851 | EPI_ISL_3801891 |
| EPI_ISL_2731451 | EPI_ISL_3539912 | EPI_ISL_3540029 | EPI_ISL_3801812 | EPI_ISL_3801852 | EPI_ISL_3801892 |
| EPI_ISL_2731452 | EPI_ISL_3539913 | EPI_ISL_3540030 | EPI_ISL_3801813 | EPI_ISL_3801853 | EPI_ISL_3801893 |
| EPI_ISL_2863898 | EPI_ISL_3539914 | EPI_ISL_3540031 | EPI_ISL_3801814 | EPI_ISL_3801854 | EPI_ISL_3801894 |
| EPI_ISL_2863899 | EPI_ISL_3539915 | EPI_ISL_3540032 | EPI_ISL_3801815 | EPI_ISL_3801855 | EPI_ISL_3801895 |
| EPI_ISL_2982735 | EPI_ISL_3539916 | EPI_ISL_3540033 | EPI_ISL_3801816 | EPI_ISL_3801856 | EPI_ISL_3801896 |
| EPI_ISL_2982743 | EPI_ISL_3539917 | EPI_ISL_3540034 | EPI_ISL_3801817 | EPI_ISL_3801857 | EPI_ISL_3801897 |
| EPI_ISL_2983373 | EPI_ISL_3539918 | EPI_ISL_3540035 | EPI_ISL_3801818 | EPI_ISL_3801858 | EPI_ISL_3801898 |
| EPI_ISL_2983452 | EPI_ISL_3539919 | EPI_ISL_3540036 | EPI_ISL_3801819 | EPI_ISL_3801859 | EPI_ISL_3801899 |
| EPI_ISL_3134853 | EPI_ISL_3539920 | EPI_ISL_3540037 | EPI_ISL_3801820 | EPI_ISL_3801860 | EPI_ISL_3801900 |
| EPI_ISL_3134854 | EPI_ISL_3539921 | EPI_ISL_3540038 | EPI_ISL_3801821 | EPI_ISL_3801861 | EPI_ISL_3801901 |
| EPI_ISL_3134855 | EPI_ISL_3539923 | EPI_ISL_3540039 | EPI_ISL_3801822 | EPI_ISL_3801862 | EPI_ISL_3801902 |
| EPI_ISL_3274747 | EPI_ISL_3539924 | EPI_ISL_3703695 | EPI_ISL_3801823 | EPI_ISL_3801863 | EPI_ISL_3801903 |
| EPI_ISL_3274750 | EPI_ISL_3540001 | EPI_ISL_3703704 | EPI_ISL_3801824 | EPI_ISL_3801864 | EPI_ISL_3801904 |
| EPI_ISL_3274764 | EPI_ISL_3540002 | EPI_ISL_3758076 | EPI_ISL_3801825 | EPI_ISL_3801865 | EPI_ISL_3801905 |
| EPI_ISL_3274790 | EPI_ISL_3540003 | EPI_ISL_3758085 | EPI_ISL_3801826 | EPI_ISL_3801866 | EPI_ISL_3801906 |
| EPI_ISL_3386143 | EPI_ISL_3540004 | EPI_ISL_3758115 | EPI_ISL_3801827 | EPI_ISL_3801867 | EPI_ISL_3801907 |
| EPI_ISL_3447445 | EPI_ISL_3540005 | EPI_ISL_3758116 | EPI_ISL_3801828 | EPI_ISL_3801868 | EPI_ISL_3801908 |
| EPI_ISL_3447452 | EPI_ISL_3540006 | EPI_ISL_3758124 | EPI_ISL_3801829 | EPI_ISL_3801869 | EPI_ISL_3801909 |
| EPI_ISL_3536204 | EPI_ISL_3540007 | EPI_ISL_3758134 | EPI_ISL_3801830 | EPI_ISL_3801870 | EPI_ISL_3801910 |
| EPI_ISL_3536331 | EPI_ISL_3540008 | EPI_ISL_3758136 | EPI_ISL_3801831 | EPI_ISL_3801871 | EPI_ISL_3801911 |
| EPI_ISL_3536483 | EPI_ISL_3540009 | EPI_ISL_3758138 | EPI_ISL_3801832 | EPI_ISL_3801872 | EPI_ISL_3801912 |
| EPI_ISL_3536492 | EPI_ISL_3540010 | EPI_ISL_3758144 | EPI_ISL_3801833 | EPI_ISL_3801873 | EPI_ISL_3801913 |
| EPI_ISL_3536501 | EPI_ISL_3540011 | EPI_ISL_3758159 | EPI_ISL_3801834 | EPI_ISL_3801874 | EPI_ISL_3801914 |
| EPI_ISL_3536502 | EPI_ISL_3540012 | EPI_ISL_3801795 | EPI_ISL_3801835 | EPI_ISL_3801875 | EPI_ISL_3801915 |
| EPI_ISL_3536503 | EPI_ISL_3540013 | EPI_ISL_3801796 | EPI_ISL_3801836 | EPI_ISL_3801876 | EPI_ISL_3801916 |
| EPI_ISL_3536504 | EPI_ISL_3540014 | EPI_ISL_3801797 | EPI_ISL_3801837 | EPI_ISL_3801877 | EPI_ISL_3801917 |
| EPI_ISL_3536512 | EPI_ISL_3540015 | EPI_ISL_3801798 | EPI_ISL_3801838 | EPI_ISL_3801878 | EPI_ISL_3801918 |
| EPI_ISL_3536519 | EPI_ISL_3540016 | EPI_ISL_3801799 | EPI_ISL_3801839 | EPI_ISL_3801879 | EPI_ISL_3801919 |
| EPI_ISL_3536522 | EPI_ISL_3540017 | EPI_ISL_3801800 | EPI_ISL_3801840 | EPI_ISL_3801880 | EPI_ISL_3801920 |
| EPI_ISL_3536527 | EPI_ISL_3540018 | EPI_ISL_3801801 | EPI_ISL_3801841 | EPI_ISL_3801881 | EPI_ISL_3801921 |
| EPI_ISL_3536531 | EPI_ISL_3540019 | EPI_ISL_3801802 | EPI_ISL_3801842 | EPI_ISL_3801882 | EPI_ISL_3801922 |
| EPI_ISL_3801923 | EPI_ISL_3803003 | EPI_ISL_3832332 | EPI_ISL_3832374 | EPI_ISL_3835281 | EPI_ISL_3922261 |
| EPI_ISL_3801924 | EPI_ISL_3825472 | EPI_ISL_3832333 | EPI_ISL_3832375 | EPI_ISL_3835287 | EPI_ISL_3944568 |

| | | | | | |
|---|---|---|---|---|---|
| EPI_ISL_3801925 | EPI_ISL_3827877 | EPI_ISL_3832334 | EPI_ISL_3832376 | EPI_ISL_3835358 | EPI_ISL_3944569 |
| EPI_ISL_3801926 | EPI_ISL_3827878 | EPI_ISL_3832335 | EPI_ISL_3832377 | EPI_ISL_3841246 | EPI_ISL_3944570 |
| EPI_ISL_3801927 | EPI_ISL_3827879 | EPI_ISL_3832336 | EPI_ISL_3832378 | EPI_ISL_3841247 | EPI_ISL_4080931 |
| EPI_ISL_3801928 | EPI_ISL_3827892 | EPI_ISL_3832337 | EPI_ISL_3832379 | EPI_ISL_3841248 | EPI_ISL_4080932 |
| EPI_ISL_3801929 | EPI_ISL_3827902 | EPI_ISL_3832338 | EPI_ISL_3832380 | EPI_ISL_3841249 | EPI_ISL_4080933 |
| EPI_ISL_3801930 | EPI_ISL_3827916 | EPI_ISL_3832339 | EPI_ISL_3832381 | EPI_ISL_3841250 | EPI_ISL_4515966 |
| EPI_ISL_3801931 | EPI_ISL_3827920 | EPI_ISL_3832340 | EPI_ISL_3832382 | EPI_ISL_3912055 | EPI_ISL_4520335 |
| EPI_ISL_3801932 | EPI_ISL_3827922 | EPI_ISL_3832341 | EPI_ISL_3832383 | EPI_ISL_3912077 | EPI_ISL_4520342 |
| EPI_ISL_3801933 | EPI_ISL_3827924 | EPI_ISL_3832342 | EPI_ISL_3832384 | EPI_ISL_3912386 | EPI_ISL_5490956 |
| EPI_ISL_3801934 | EPI_ISL_3827926 | EPI_ISL_3832343 | EPI_ISL_3832385 | EPI_ISL_3912389 | EPI_ISL_5490974 |
| EPI_ISL_3801935 | EPI_ISL_3827929 | EPI_ISL_3832344 | EPI_ISL_3832386 | EPI_ISL_3912391 | EPI_ISL_5491028 |
| EPI_ISL_3801936 | EPI_ISL_3827930 | EPI_ISL_3832345 | EPI_ISL_3832387 | EPI_ISL_3912392 | EPI_ISL_5491035 |
| EPI_ISL_3801937 | EPI_ISL_3827933 | EPI_ISL_3832346 | EPI_ISL_3832388 | EPI_ISL_3912393 | EPI_ISL_5491128 |
| EPI_ISL_3801938 | EPI_ISL_3827935 | EPI_ISL_3832347 | EPI_ISL_3832389 | EPI_ISL_3912394 | EPI_ISL_5491139 |
| EPI_ISL_3802976 | EPI_ISL_3827936 | EPI_ISL_3832348 | EPI_ISL_3832390 | EPI_ISL_3912395 | EPI_ISL_6100863 |
| EPI_ISL_3802977 | EPI_ISL_3827939 | EPI_ISL_3832349 | EPI_ISL_3832391 | EPI_ISL_3912396 | EPI_ISL_6100864 |
| EPI_ISL_3802978 | EPI_ISL_3827940 | EPI_ISL_3832350 | EPI_ISL_3832392 | EPI_ISL_3912397 | EPI_ISL_6100865 |
| EPI_ISL_3802979 | EPI_ISL_3827943 | EPI_ISL_3832351 | EPI_ISL_3832393 | EPI_ISL_3912398 | EPI_ISL_6100866 |
| EPI_ISL_3802980 | EPI_ISL_3827944 | EPI_ISL_3832352 | EPI_ISL_3832394 | EPI_ISL_3912399 | EPI_ISL_6100867 |
| EPI_ISL_3802981 | EPI_ISL_3827945 | EPI_ISL_3832353 | EPI_ISL_3832447 | EPI_ISL_3912400 | EPI_ISL_6100868 |
| EPI_ISL_3802982 | EPI_ISL_3827946 | EPI_ISL_3832354 | EPI_ISL_3832448 | EPI_ISL_3912401 | EPI_ISL_6100869 |
| EPI_ISL_3802983 | EPI_ISL_3827948 | EPI_ISL_3832355 | EPI_ISL_3832449 | EPI_ISL_3912402 | EPI_ISL_6100870 |
| EPI_ISL_3802984 | EPI_ISL_3827949 | EPI_ISL_3832356 | EPI_ISL_3832450 | EPI_ISL_3912403 | EPI_ISL_6100871 |
| EPI_ISL_3802985 | EPI_ISL_3827950 | EPI_ISL_3832357 | EPI_ISL_3832451 | EPI_ISL_3912404 | EPI_ISL_6100876 |
| EPI_ISL_3802986 | EPI_ISL_3827951 | EPI_ISL_3832358 | EPI_ISL_3832452 | EPI_ISL_3912406 | EPI_ISL_6100877 |
| EPI_ISL_3802987 | EPI_ISL_3827953 | EPI_ISL_3832359 | EPI_ISL_3832453 | EPI_ISL_3914048 | EPI_ISL_6100878 |
| EPI_ISL_3802988 | EPI_ISL_3827954 | EPI_ISL_3832360 | EPI_ISL_3832454 | EPI_ISL_3914214 | EPI_ISL_6100879 |
| EPI_ISL_3802989 | EPI_ISL_3827955 | EPI_ISL_3832361 | EPI_ISL_3832460 | EPI_ISL_3914313 | EPI_ISL_6100880 |
| EPI_ISL_3802990 | EPI_ISL_3827957 | EPI_ISL_3832362 | EPI_ISL_3832462 | EPI_ISL_3922205 | EPI_ISL_6100881 |
| EPI_ISL_3802991 | EPI_ISL_3827958 | EPI_ISL_3832363 | EPI_ISL_3832464 | EPI_ISL_3922214 | EPI_ISL_6100882 |
| EPI_ISL_3802992 | EPI_ISL_3827963 | EPI_ISL_3832364 | EPI_ISL_3832465 | EPI_ISL_3922218 | |
| EPI_ISL_3802993 | EPI_ISL_3827969 | EPI_ISL_3832365 | EPI_ISL_3832466 | EPI_ISL_3922221 | |
| EPI_ISL_3802994 | EPI_ISL_3827970 | EPI_ISL_3832366 | EPI_ISL_3832469 | EPI_ISL_3922225 | |
| EPI_ISL_3802995 | EPI_ISL_3827971 | EPI_ISL_3832367 | EPI_ISL_3832470 | EPI_ISL_3922231 | |
| EPI_ISL_3802996 | EPI_ISL_3827976 | EPI_ISL_3832368 | EPI_ISL_3832472 | EPI_ISL_3922234 | |
| EPI_ISL_3802997 | EPI_ISL_3827987 | EPI_ISL_3832369 | EPI_ISL_3832473 | EPI_ISL_3922237 | |
| EPI_ISL_3802998 | EPI_ISL_3828020 | EPI_ISL_3832370 | EPI_ISL_3835245 | EPI_ISL_3922242 | |
| EPI_ISL_3802999 | EPI_ISL_3832329 | EPI_ISL_3832371 | EPI_ISL_3835250 | EPI_ISL_3922252 | |
| EPI_ISL_3803001 | EPI_ISL_3832330 | EPI_ISL_3832372 | EPI_ISL_3835275 | EPI_ISL_3922258 | |
| EPI_ISL_3803002 | EPI_ISL_3832331 | EPI_ISL_3832373 | EPI_ISL_3835279 | EPI_ISL_3922260 | |
